# Temporal Trends in Thromboembolic Event Hospitalizations in Patients with Atrial Fibrillation in the United States

**DOI:** 10.1101/2024.07.03.24309193

**Authors:** Moises A. Vasquez, Crystal Lihong Yan, Litsa K. Lambrakos, Alex Velasquez, Jeffrey J. Goldberger, Raul D. Mitrani

**Affiliations:** Division of Internal Medicine, University of Miami Miller School of Medicine/Jackson Health System, Miami, Florida, USA; Division of Cardiovascular Medicine, University of Miami Health System, Miami, Florida, USA

**Author notes:** Corresponding author:, Address: 1611 NW 12th Ave, C-600D, Miami, FL 33136, Department of Internal Medicine, University of Miami Miller School of Medicine/Jackson Memorial Hospital, Miami, Florida, USA. Raul D. Mitrani was supported in part by the Palm Health Foundation & Marmot Foundation. **Abbreviations:** AF: Atrial Fibrillation; AC: Anticoagulation; LAAC: Left Atrial Appendage Occlusion; AIS: Acute ischemic stroke; SEE: Systemic Embolic Events; TIA: Transient Ischemic Attack.

**Keywords:** Atrial Fibrillation, Temporal Trends, Acute Ischemic Stroke, Transient Ischemic Attack, Systemic Embolic Events, Oral Anticoagulation, Direct Oral Anticoagulants, Vitamin-K Antagonists, Non-Vitamin K Antagonists, Left Atrial Appendage Occlusion

## Abstract

**Background:** Direct oral anticoagulants and percutaneous left atrial appendage occlusion (LAAC) devices were approved for use in 2010 and 2015, respectively. It is unknown to what extent, if any, these new stroke preventive therapies have impacted hospitalizations for thromboembolic (TE) events in the United States.

**Methods:** We used the National Inpatient Sample database to describe trends over time in AF-related hospitalizations for acute ischemic stroke (AIS), transient ischemic attacks (TIA), and systemic embolic events (SEE) between 2010 and 2021. Trends in anticoagulation use (AC) and percutaneous LAAC procedures among hospitalized patients with AF were also analyzed.

**Results:** A total of 1,692,373 hospitalizations for TE were identified between 2010 and 2021: 798,413 from 2010 to 2015 (mean age 78.9 [standard deviation (SD) 10.7] years; 57.8% female) and 893,960 from 2016 to 2021 (mean age 77.5 [SD 10.9] years; 53.1% female). There was a significant increase in rates of AC use (21.2% of inpatients with AF in 2010 vs 42.4% of inpatients with AF in 2021; P_trend_ <0.001) and rates of percutaneous LAAC procedures (0.1/100,000 US adults vs to 17.7/100,000 US adults P_trend_ <0.001) among inpatients with AF. Between 2010 to 2021, age-adjusted AIS hospitalizations increased (30.8 hospitalizations /100,000 US adults vs 35.9 hospitalizations/ 100,000 US adults; P_trend_ <0.001) and peaked in 2019. Age-adjusted SEE hospitalizations (2.5 hospitalizations /100,000 US adults vs 1.3 hospitalizations/ 100,000 US adults; P_trend_ <0.001) and TIA hospitalization(8.5 hospitalizations /100,000 US adults vs 4.3 hospitalizations/ 100,000 US adults; P_trend_ <0.001) peaked in 2011 and then decreased. Between 2016 and 2021, age-adjusted AIS hospitalizations plateaued (37.7 hospitalizations/100,000 US adults vs 35.9 hospitalizations/100,000 US adults; APC −4.7%; P_trend_ = 0.197). The total of all TE hospitalizations remained stable (41.8 hospitalizations/100,000 US adults vs 41.5 hospitalizations/100,000 US adults; P_trend_ 0.689).

**Conclusion:** Despite increased anticoagulant and the initial use of percutaneous LAAO, AF-associated TE event hospitalizations in the US have remained stable from 2010 to 2021. Hospitalizations for AF-related TIA and SEE consistently decreased from 2010 to 2021 while AF-related AIS hospitalizations increased from 2010 to 2015 and then plateaued from 2016 to 2021.

## Introduction

Atrial fibrillation (AF) is associated with a 4-to 5-fold increased risk of stroke and a 3-fold increased risk of extracranial systemic embolic events (SEE) (1,2). It accounts for 25% of cerebrovascular accidents in the United States (US) (2,3). Although vitamin K antagonists (VKA) greatly reduce the risk of stroke in AF, compliance with AC therapy had been suboptimal, in part due to low adherence and potential or realized bleeding complications (4,5). Following the introduction of direct oral anticoagulants (DOAC), first approved in 2010 by the US Food and Drug Administration (FDA), these medications have emerged as the primary therapeutic alternative to VKAs. In 2015, regulatory approval of the first transcatheter left atrial appendage closure (LAAC) device introduced an alternative nonpharmacologic stroke prevention therapy for patients who are unable to tolerate or have other justification to avoid long-term anticoagulation (AC) (6,7). However, whether these milestones have impacted hospitalizations for AF-related thromboembolic (TE) events in the US remains unclear.

Previous National Inpatient Sample (NIS) database analyses had reported increasing prevalence of AF in acute ischemic stroke (AIS) and transient ischemic attacks (TIA) hospitalizations prior to 2010 (8,9). Another NIS analysis had reported a rise in AIS hospitalizations among patients with AF over 90 years of age from 2004 to 2015 (10). More recent data from outside the US have described changes in AC use and ischemic stroke rates among patients with AF over time, consistently reporting a decline in AF-related AIS from 2010 to 2020, paralleling an increase in the use of anticoagulation therapy (11–16). A more recent NIS study reported decreasing prevalence of comorbid AF in patients admitted for acute ischemic stroke (17). These studies have not focused on TIAs and SEEs. It is unknown whether the advancements in stroke prevention therapies for patients with AF have impacted trends in hospitalizations for AIS and other TE events in the US. To test this hypothesis, we used the NIS database to analyze temporal trends in hospitalizations for AF-associated AIS, TIAs, and extracranial SEEs in the US from 2010 to 2021.

## Methods

### Study Data

We conducted a retrospective study using the NIS database, the largest inpatient care database in the US, which is derived from the billing data submitted by hospitals to statewide data organizations as part of the Healthcare Cost and Utilization Project (HCUP) (18). Each observation in the NIS represents an individual hospitalization with a primary diagnosis, up to 39 secondary diagnoses, and up to 25 procedure codes. To estimate the total number of hospitalized patients across the United States, a uniform sampling and weighting approach recommended by HCUP was utilized. The NIS has been used extensively to assess national trends and inpatient outcomes in patients with AF, among other diseases (17,19–22).

IRB approval exemption was granted as a deidentified administrative database was used. We adhered strictly to the NIS survey methodology on data interpretation, research design, and data analysis as described by the HCUP. Guidelines according to the Strengthening the Reporting of Observational Studies in Epidemiology Statement were followed, along with methodologies for best practices using claims datasets (23).

### Study Population

Adult hospitalizations with primary diagnosis codes for TE events (AIS, TIA, and SEE) and concurrent secondary codes for AF from 2010 to 2021 were identified using the International Classification of Diseases, Ninth Revision, Clinical Modification (ICD-9-CM) and International Statistical Classification of Diseases and Related Health Problems, Tenth Revision (ICD-10) codes. ICD-9-CM codes were used from 2010 to the third quarter of 2015, and ICD-10 codes were used from the fourth quarter of 2015 to 2021. Records with ICD codes for rheumatic mitral valve stenosis or the presence of a mitral prosthetic valve were excluded from the study. Data were divided into two temporal periods: the first included hospitalizations from 2010 to 2015, and the second included hospitalizations from 2016 to 2021. Baseline patient characteristics and clinically relevant comorbidities were included, using previously validated ICD-10 and ICD-9 codes, when available. Components of the CHA2DS2-VASc score were defined by the age of 65 to 74 years, age ≥75 years, a diagnosis of heart failure, hypertension, diabetes mellitus, previous ischemic stroke, female sex, and vascular disease (prior myocardial infarction, prior revascularization, peripheral arterial disease, or coronary artery disease). The ICD-10 and ICD-9-CM codes used to identify comorbidities are provided in Supplemental Table 1.

### Outcome definition

To identify TE events, we exclusively utilized the primary hospitalization diagnosis to increase the probability that the hospitalization was attributable to one of the adverse events identified in our study rather than a subsequent complication or a previously documented issue. SEE included all cases of presumed embolic acute limb ischemia or acute visceral ischemia. Hospitalization rates were presented as a rate per 100,000 US adult population. This was calculated by dividing total weighted number of TE hospitalizations with a diagnosis of AF for a specific year (numerator), by the US census adult population estimate for that year (denominator) then multiplied by 100,000 (24). These hospitalizations were further age adjusted based on the US standard population from the year 2000.

Temporal changes in the use of stroke prevention strategies (long-term AC use and percutaneous LAAC procedures) among all hospitalizations with primary or secondary diagnoses codes for AF were also described. Changes in frequency of the ICD-9 code V58.69 and ICD-10 code Z79.01 for current AC use among all hospitalized patients with a diagnosis of AF (primary or secondary) was used to determine trends in AC use. Yearly AC use rates were calculated by dividing the weighted number of patients with a diagnosis of AF and concurrent use of AC (numerator) by the total weighted number of patients with AF hospitalized for any diagnosis (denominator) in the NIS. To calculate annual percutaneous LAAC procedure rates, the numerator was substituted with the weighted number of inpatients with AF with a procedure code for percutaneous LAAC placement. Annual percent changes (APC) between years were described.

### Statistical Analysis

Survey methodology was used for all analyses to account for the clustering and stratification of hospitalizations in the NIS as recommended by the Agency for Healthcare Research and Quality (25). The trend weight files were merged onto the original NIS files by year and hospital identification number. For years before 2012, the trend weight was used to create national estimates for trend analysis. For 2012 and after, no trend weight was needed, and the regular discharge weight was used, consistent with the redesigned NIS trend analysis (26). Cochran-Mantel-Haenszel trend test was used for categorical variables, and linear regression was used for continuous variables. Continuous variables were expressed as means and standard deviations. Categorical data were expressed as frequencies. Categorical variables were compared between cohorts using Pearson’s Chi-Squared (χ2) test. All reported *P* values were two-sided, and the significance cut-off was set at 0.05. The Bonferroni adjustment was used to adjust for multiple testing between categorical ^2^ tests. Continuous variables were compared using the independent samples Student t-test if normally distributed, and Mann-Whitney U test for non-normally distributed continuous variables. All statistical analyses were performed using SPSS (IBM SPSS Statistics, Version 28.0, IBM Corporation, Armonk, NY).

## Results

A total of 1,692,373 hospitalizations for AF-related TE events were identified between 2010 and 2021. Temporal changes in baseline characteristics are listed in Table 1. There were 798,413 hospitalizations (47.2%) from 2010 to 2015 (mean age 78.9 [standard deviation (SD) 10.7] years; 57.8% female) and 893,960 hospitalizations (52.8%) from 2016 to 2021 (mean age 77.5 [SD 10.9] years; 53.1% female). Hospitalizations for AIS accounted for 75.9% of all TE hospitalizations in 2010 to 2015 and 84.2% in 2016 to 2021 (P <0.001). A significant increase in the prevalence of stroke risk factors between periods was observed, such as hypertension (88.9% vs. 83.8%; P <0.001), hyperlipidemia (61.3% vs. 52.1%; P <0.001), diabetes (36.0% vs. 31.7%; P <0.001), and tobacco use (32.6% vs 21.1%; P <0.001). Inpatients in the 2016-2021 period more frequently had a history of previous stroke or TIA (16.3% vs 14.5%; P <0.001) compared to 2010-2015. The mean CHA2DS2-VASc score increased from 4.4 in 2010 to 2015 to 4.5 in 2016 to 2021 (P <0.001). The proportion of inpatients with a CHA2DS2-VASc of 2 to 4 decreased (48.9% vs 51.4%, P <0.001) but the proportion of inpatients with a CHA2DS2-VASc ≥ 5 increased (47.6% vs 45.2%, P <0.001).

### Thromboembolic events

Between 2010 and 2021, the total of all AF-associated TE hospitalizations had a non-significant increase from 50.9 hospitalizations/100,000 US adults to 55.6 hospitalizations /100,00 US adults (APC +9.2%; P_trend_ 0.148), peaking at 61.1 hospitalizations/100,000 US adults in 2017. From 2017 to 2021, crude hospitalizations for TE events remained stable (61.1 hospitalizations/100,000 US adults vs 55.7 hospitalizations/100,000 US adults; P_trend_ = 0.0967). After age-adjusting, the total of all TE hospitalizations also remained stable from 2010 to 2021 (41.8 hospitalizations/100,000 US adults vs 41.5 hospitalizations/100,000 US adults; APC - 0.7%; P_trend_ = 0.689), with a peak occurring in 2017 at 46.9 hospitalizations/100,000 US adults. From 2017 to 2021, age-adjusted hospitalizations TE events also remained stable (46.9 hospitalizations/100,000 US adults vs 41.5 hospitalizations/100,000 US adults; P_trend_ = 0.061).

### Ischemic Stroke

The temporal changes in crude and age-adjusted hospitalizations for AF-associated AIS are shown in Figure 2A and Figure 3A, respectively. Crude AIS hospitalizations increased from 37.5 hospitalizations /100,000 US adults to a peak of 51.7 hospitalizations/100,000 US adults in 2019, before declining to 48.1 hospitalizations /100,000 US adults in 2021 (APC +28.2%; P_trend_ <0.001). Age-adjusted AIS hospitalizations increased from 30.8 hospitalizations /100,000 US adults in 2010 to 35.9 hospitalizations/ 100,000 US adults in 2021 (APC +16.5%; P_trend_ <0.001), reaching a 12-year high of 39.1 hospitalizations/ 100,000 in 2019. From 2016 to 2019, age-adjusted AIS hospitalizations plateaued (37.7 hospitalizations /100,000 US adults vs 39.1 hospitalizations /100,000 US adults; P=0.064), before decreasing in 2020 to 34.5 hospitalizations/100,000 US adults.

**Figure 1.**
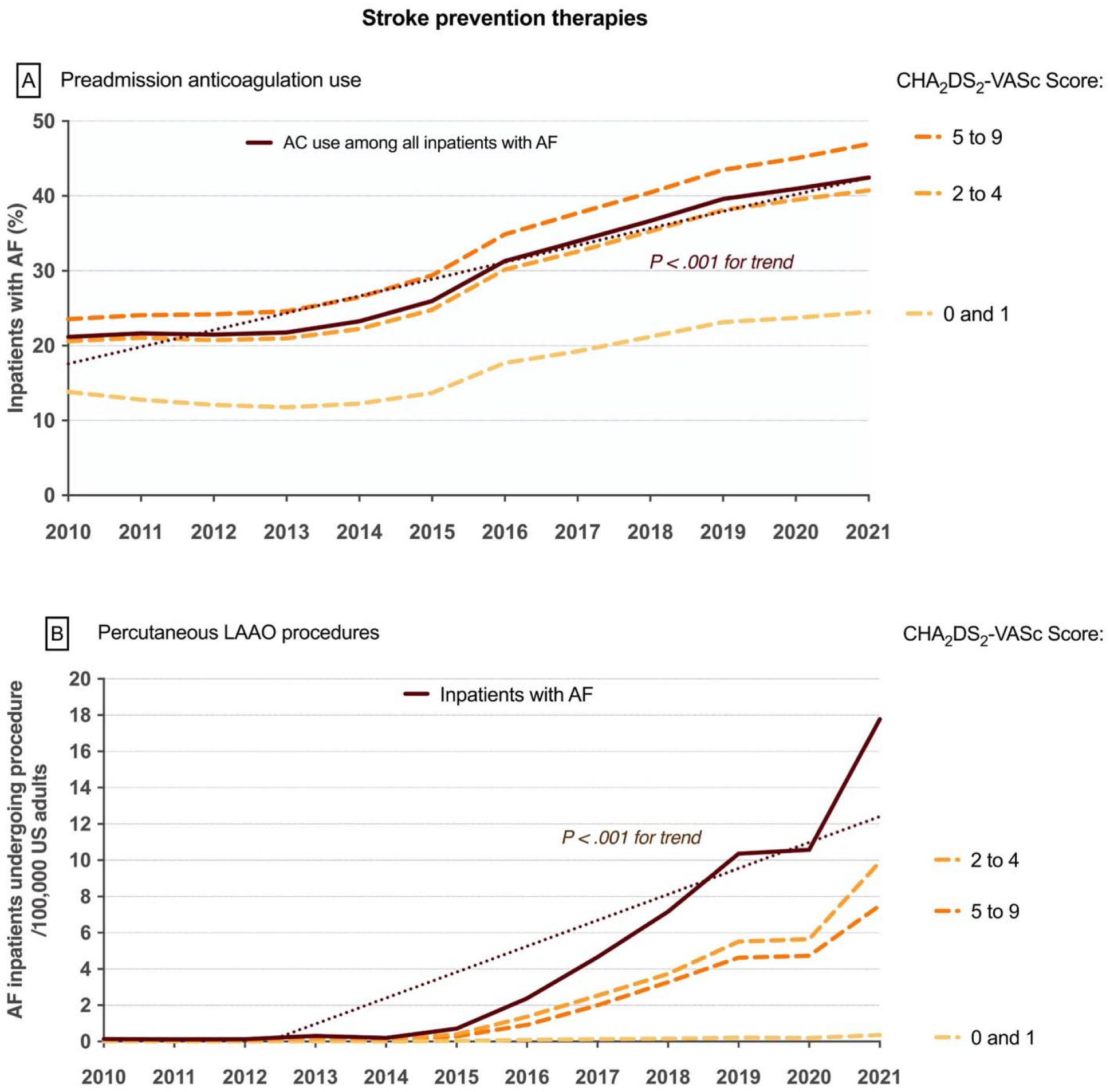
Temporal trends in anticoagulation use (A) and transcatheter LAAC procedures (B) among patients hospitalized with atrial fibrillation, stratified by CHA2DS2-VASc score. Dotted line represents mean trend and solid line represents year to year trend. Colored dashed lines represent CHA2DS2-VASc score categories. *LAAC = Left Atrial Appendage Occlusion*

**Figure 2.**
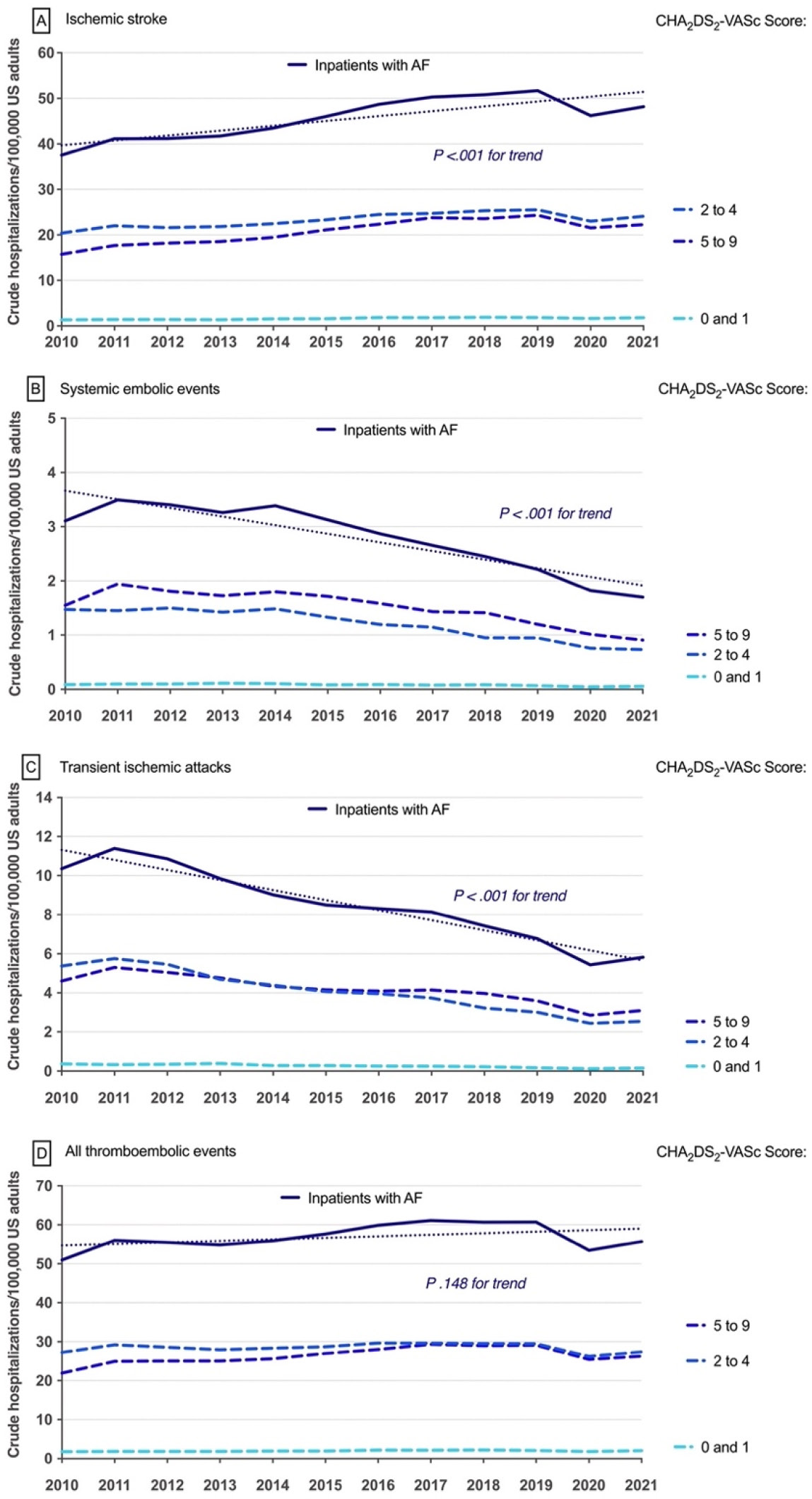
Temporal trends in crude hospitalizations for ischemic stroke (A), SEE (B), TIAs (C), and the composite outcome (D) among patients with atrial fibrillation, stratified by CHA2DS2-VASc score. Dotted line represents mean trend and solid line represents year to year trend. Colored dashed lines represent CHA2DS2-VASc score categories.

**Figure 3.**
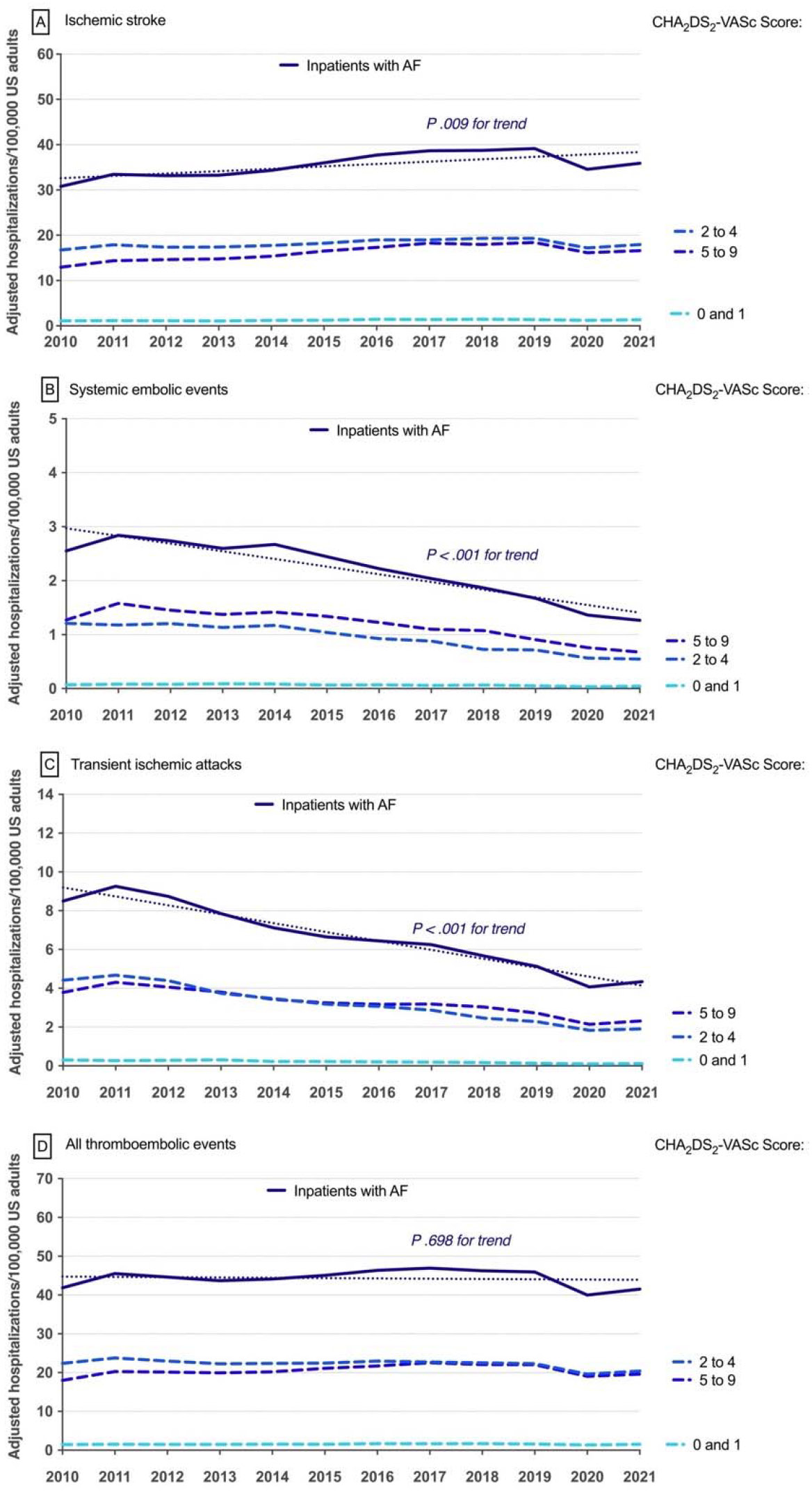
Temporal trends in age-adjusted hospitalizations for ischemic stroke (A), SEE (B), TIAs (C), and all TE event hospitalizations (D) among patients with atrial fibrillation, stratified by CHA2DS2-VASc score. Dotted line represents mean trend and solid line represents year to year trend. Colored dashed lines represent CHA2DS2-VASc score categories.

**Figure 4.**
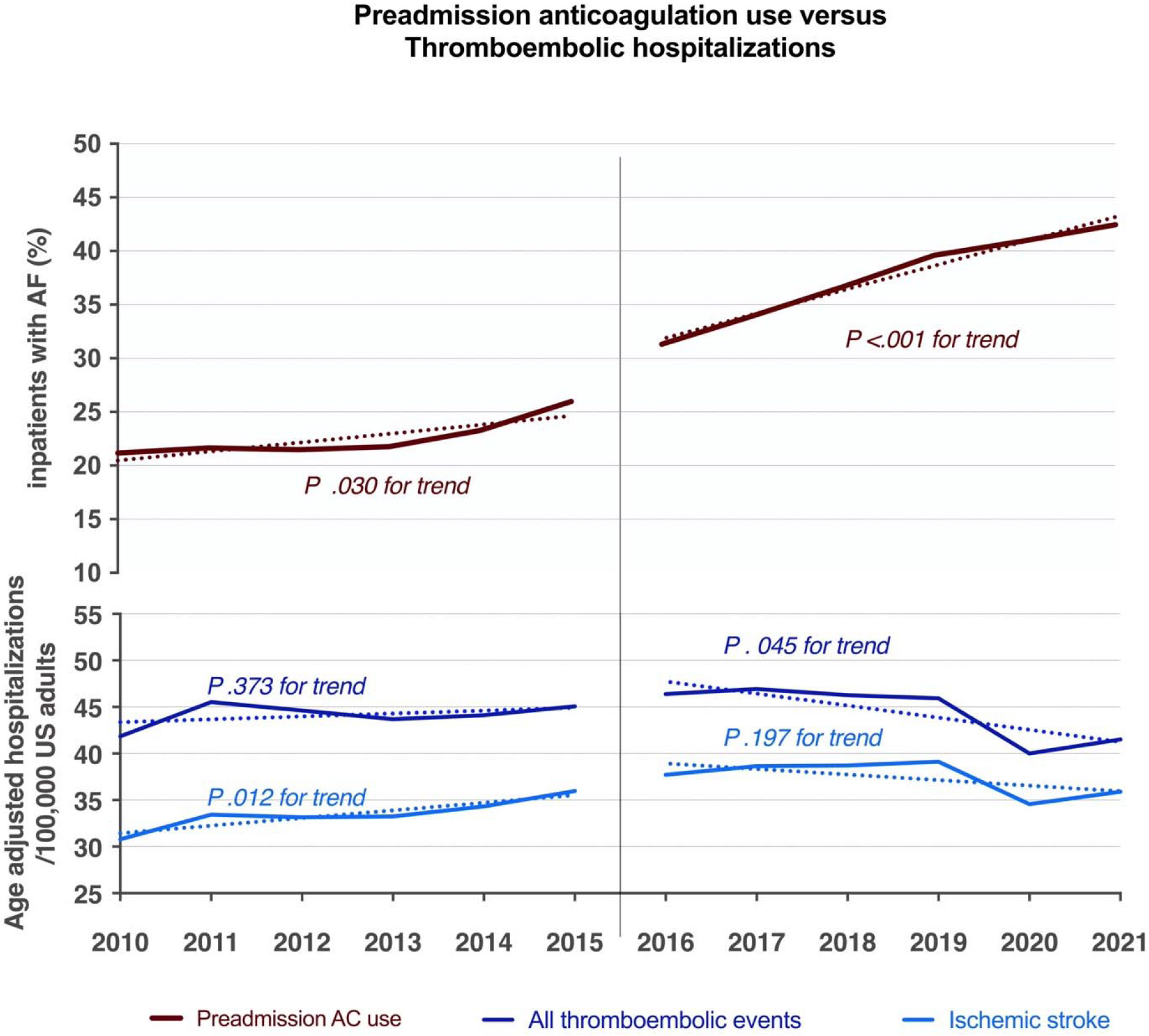
Comparison of temporal trends in preadmission anticoagulation use, in contrast to hospitalizations for ischemic stroke and all thromboembolic events in patients with atrial fibrillation. Dotted line represents mean trend and solid line represents year to year trend. Note the difference in scale in Y axis.

### Systemic embolic events

The temporal changes in crude and age-adjusted hospitalizations for AF-associated SEEs are shown in Figure 2B and Figure 3B respectively. Crude SEE hospitalizations decreased from 3.1 hospitalizations/100,000 US adults to 1.7 hospitalizations/100,000 US adults between 2010 and 2021 (APC −45.1%; P_trend_ <0.001). Age-adjusted SEE hospitalizations decreased from 2.5 hospitalizations /100,000 US adults in 2010 to 1.3 hospitalizations/ 100,000 US adults in 2021 (APC −48.0%; P_trend_ <0.001).

### Transient Ischemic Attacks

The temporal changes in crude and age-adjusted hospitalizations for AF-associated TIAs are shown in Figure 2C and Figure 3C respectively. Crude TIA hospitalizations increased from 10.3 hospitalizations /100,000 US adults in 2010 to a peak of 11.4 hospitalizations /100,000 US adults in 2011, then declined to 5.8 hospitalizations/100,000 US adults by 2021 (APC −43.6%; P_trend_ <0.001). Age-adjusted TIA hospitalizations also decreased from 8.5 hospitalizations /100,000 US adults in 2010 to 4.3 hospitalizations/ 100,000 US adults in 2021 (APC −49.4%; P_trend_ <0.001).

### Comparison between periods

Age-adjusted trend comparisons between the two periods are shown in Figure 3. Between 2010 and 2015, age-adjusted AIS hospitalizations increased from 30.7 hospitalizations/100,000 US adults to 35.9 hospitalizations/100,000 US adults (APC +16.9%; P_trend_ = 0.012). Between 2016 and 2021, no significant trend was observed in age-adjusted AIS hospitalizations (37.7 hospitalizations/100,000 US adults vs 35.9 hospitalizations/100,000 US adults; APC −4.7%; P_trend_ = 0.197). Between 2010 to 2015, the total of all age-adjusted TE hospitalizations was unchanged (41.8 hospitalizations/100,000 US adults vs 45.0 hospitalizations/100,000 US adults; APC +7.6%; P_trend_ = 0.373). However, between 2016 to 2021 TE hospitalizations significantly decreased from 46.3 hospitalizations/100,000 US adults to 41.5 hospitalizations/100,000 US adults (APC −10.3%; P_trend_ = 0.045).

The number of inpatients with AF on long-term AC increased in both the 2010-2015 period (292/100,000 US adults vs 410/100,000 US adults; APC +40.4%; P_trend_ = 0.022) and the 2016-2021 period (505 inpatients/100,000 US adults vs 705 inpatients/100,000 US adults; APC +39.6%; P_trend_ = 0.013).

### Stroke prevention therapies

Temporal trends in AC use and percutaneous LAAC procedures stratified by CHA2DS2-VASc score are shown in Figure 1. Out of all AF hospitalizations between 2010 and 2021, the proportion of patients on documented AC therapy increased from 21.2% in 2010 to 42.4% in 2021 (P_trend_ <0.001). with a proportional increase among all CHA2DS2-VASc groups. Annual trends in AC use stratified by CHA2DS2-VASc are shown in Figure 1A.

The number of LAAC procedures on patients hospitalized with AF also significantly increased from 0.1 procedures/100,000 US adults in 2010 to 17.7 procedures/100,000 US adults in 2021 (APC +17,600%; P_trend_ <0.001). These procedures had the largest increase in patients with CHA2DS2-VASc scores between 2 and 4 (0.11 procedures/100,000 US adults vs 9.9 procedures /100,000 US adults; APC +8,900%; P_trend_ <0.001), and ≥5 (0.02 procedures/100,000 US adults vs 7.5 hospitalizations/100,000 US adults; APC +37,400%; P_trend_ <0.001). By 2019, a total of 66,171 percutaneous LAAC procedures were identified. This number increased to 139,546 by 2021. Yearly frequency of LAAO procedures among all AF inpatients increased from 0.03% in 2015 to 1.2% in 2021 (Ptrend <0.001).

## Discussion

In this analysis of over a million hospitalizations in patients with AF-associated TE events, the key findings are: 1) Between 2010 and 2021, the use of AC therapy among patients with AF has significantly increased. The rate of transcatheter LAAC procedures also increased dramatically from 2016 to 2021. 2) Despite the increased use of stroke prevention therapies in the hospitalized AF population, there was no substantial reduction in the AF-related hospitalizations for TE events from 2010 to 2021. However, AF-related TE event hospitalizations significantly decreased from 2016 to 2021. Hospitalizations for AIS increased from 2010 to 2015, and plateaued from 2016 onwards; therefore, while the rise in AC use and percutaneous LAAC procedures was not associated with a decline in AIS, the rate is no longer rising.

Previous NIS analyses had reported a similar uptick in AIS hospitalizations in the general population between 2009 to 2018 and among patients with AF over 90 years of age from 2004 to 2015 (10,21). Most recently, a NIS study looking at the prevalence of AF in hospitalizations for acute IS in the United States, found that AF prevalence increased significantly over the period 2010– 2020 but from 2015-2020, AF prevalence declined in womenl7l60years and plateaued in in menl7l60years (17). This aligns with cohort studies from China and Denmark noting a rise in AF-associated ischemic events into the early 2010s (27,28). The authors suggested this increase may indicate a true increase in stroke incidence or may represent surveillance bias due to better stroke diagnosis (10,21). Our study notes a small but significant increase in AF-associated AIS hospitalizations from 2010 to 2015, followed by a plateau from 2016 to 2021. Annual rates of all AF-associated TE events (including AISs, SEEs, and TIAs) peaked in 2017 and remained stable to 2021. This shift may be attributed to the increased use of AC noted in our study, consistent with previous reports from high-income countries that AF-related stroke rates are declining with increased availability of DOACs (11–17).

Our data also spans the COVID-19 pandemic years (2020 and 2021), which saw a marked decline in AF-associated TE hospitalizations in our study and in previous studies (29–31). While rising AC use may explain the declining TE hospitalization trends up to 2019, it is likely that the abrupt downturn in AF-related TE hospitalizations seen in 2020 was influenced by pandemic-related factors. Factors that may explain the decrease in hospitalizations during the pandemic include patient fear of COVID-19 exposure, lockdown measures, and decreased third-party detection of acute stroke symptoms (29,30). However, COVID itself could have been a causative factor for stroke in patients with AF (32). Therefore, the true impact of antithrombotic therapies on strokes and TE events in 2020 and beyond is difficult to discern. As these measures loosened in 2021, a small uptick in TE event hospitalizations was seen.

The proportion of inpatients with AF on AC therapy doubled from 2010 to 2021, with 41.2% of inpatients reported on AC by 2021. Although not indicative of the true rates of AC use among all patients with AF in the US, this certainly indicates a considerable rise in AC use in this population. This increase is supported by data from other high-income countries showing a rise in AC use among patients with AF after the launch of DOACs and their incorporation into guidelines as the first line AC therapy (11–16). DOACs offer advantages in terms of safety, efficacy, and convenience over VKAs, facilitating adherence. Our findings particularly highlight a marked increase after 2015. The 2014 shift to the CHA2DS2-VASc scoring from CHADS2 in the AHA/ACC/HRS AF guidelines could have played a role in this rise by broadening the indications for AC prescription (33–35). Nevertheless, it is concerning that despite this increase, the trend in hospitalizations for AIS did not decrease. In contrast, hospitalizations for TIAs consistently decreased starting in 2011. This discordance is partially explained by the change in the definition of TIA in 2009 from symptoms to imaging results (i.e., absence of stroke findings on MRI) and is consistent with the observed decline in TIA hospitalizations beginning in 2011 and an increase in AIS around the same time. It is possible that increases in AIS were also driven by increased stroke detection using advanced imaging, such as MRI and other factors such as a rise in AF prevalence and increased strokes in previously undetected (and thus untreated) AF. These confounding factors may have obscured the impact of DOACs and LAAC on patients presenting with stroke symptoms.

While AC usage has grown, a notable portion of inpatients with AF were not on AC by 2021. Patients presenting with new onset AF, underutilization of the ICD code for AC use, and the increasing number of patients with LAAC procedures may have also impacted the reported proportion of inpatients with AF on AC therapy in our data. Many patients also have a contraindication to AC, such as bleeding and fall risk. The utilization of LAAC in patients at high risk for stroke who are not suitable candidates for AC could potentially lead to a decrease in ischemic events in this population in the future. Our analysis shows that transcatheter LAAC procedures exponentially increased in 2015, following FDA approval of the first percutaneous endovascular LAAC device (38, 39). This rise plateaued in 2020 amid the pandemic, consistent with reported decreases in other transcatheter cardiac procedures, but surged in 2021, possibly due to relaxed pandemic policies and a new device FDA approval (40,41). Given the retrospective nature of our data, it is difficult to determine the impact this rise may have had in reducing TE events in patients with AF by 2021. Several randomized controlled trials are currently ongoing, looking to compare LAAC to DOACs in patients with AF without a contraindication for AC, such as the CHAMPION-AF (NCT 04394546) and the CATALYST (NCT 04226547) trials. These trials aim to show non-inferiority for stroke prevention but superiority for bleeding, which would make LAAC an attractive therapy over the lifelong need for AC.

### Limitations

There are several limitations to this study. The data sourced from the NIS database utilizes ICD codes, which are used for billing purposes and to encapsulate clinical scenarios retrospectively. This reliance on “claims data” does not provide insights into clinical presentations. Moreover, the accuracy and consistency of coding within the NIS might suffer due to individual provider preferences and variations in clinical documentation. In this regard, the rates of anticoagulant use might be underrepresented if the prescriptions were not adequately documented, as previously described rates in the US are around 55-65% (16). Thus, our report on trends in long-term anticoagulant use should be viewed with caution as we are unable to ascertain the validity of these codes. The database does not disclose specific details regarding the anticoagulants prescribed. The NIS focuses on hospital discharge data rather than individual patient records, making it uncertain how much data from patients with repeated hospitalizations influenced the analysis. It may also omit certain confounding variables not recorded within its scope, such as patient preferences and the reasoning behind healthcare providers’ decisions on AC prescriptions. Although all TE hospitalizations had a concurrent secondary code for AF, we are unable to say whether AF was causally related to these TE events. AF may coexist in patients with other TE event etiologies. Due to limitations of our administrative claims data, we are unable to differentiate if AF was present before hospitalizations or was detected after the TE event.

### Conclusion

AF-associated TE event hospitalizations in the US have remained stable from 2010 to 2021. Hospitalizations for AF-associated TIAs and SEEs have substantially decreased between 2010 and 2021. Hospitalizations for AIS increased from 2010-2016 and then plateaued from 2016 to 2021. As more patients are placed on AC therapy or receive percutaneous LAAC procedures, patient-level data is needed to determine the impact on hospitalizations for TE events, particularly AIS.

## Data Availability

All data produced are available online at https://hcup-us.ahrq.gov/nisoverview.jsp

## Notes

### Competing Interest Statement

The authors have declared no competing interest.

### Funding Statement

This study did not receive any funding.

