## Supplemental files for "Temporal Trends in Thromboembolic Event Hospitalizations in Patients with Atrial Fibrillation in the United States"

**Supplemental Table 1.** ICD-10 diagnosis (CM) and procedure (PCS) codes

| **Inclusion** | **ICD-10-CM/PCS codes** | **ICD-9 codes** |
| --- | --- | --- |
| Atrial Fibrillation | I480, I481, I4811, I4819, I482, I4820, I4821, I4891 | 42731 |

| **Stroke Prevention therapies** | **ICD-10-CM/PCS codes** | **ICD-9 codes** |
| --- | --- | --- |
| Long-term AC | Z7901 | V5861 |
| LAAC procedures | 02573ZK, 02574ZK, 02B73ZK,02B74ZK, 02L73CK, 02L73DK, 02L73ZK, 02L74CK, 02L74DK, 02L74ZK | 3790 |

| **Baseline characteristics** | **ICD-10 CM codes** | **ICD-9 codes** |
| --- | --- | --- |
| **Comorbidities** | | |
| Diabetes mellitus | E10.x, E11.x, E12.x E13.x | 250.0-250.3, 250.4-250.9 |
| Hypertension | I10.x, I11.x-I13.x, I15.x | 401.x 402.x-405.x |
| Tobacco use | F17.x, Z72.0, Z87.891, Z7722 | 64900, 64901, 64902, 64903, 64904, 98984, V1582, 3051 |
| Alcohol abuse | F10, Z72.1 | 265.2, 291.1-291.3, 291.5-291.9, 303.0, 303.9, 305.0, 357.5, 425.5, 535.3, 571.0-571.3, 980.x, V11.3 |
| Obesity | E66.x, Z683x, Z684x, Z6854 | 278.0 |
| Coronary artery disease | I25.x | 4140x, , 4142, 4143, 4144, 4148, 4149 |
| Hyperlipidemia | E78.x | 272x |
| Peripheral vascular disease | I70.x, I71.x, I73.1, I73.8, I73.9, I77.1, I79.0, I79.2, K55.1, K55.8, K55.9, Z95.8, Z95.9 | 093.0, 437.3, 440.x, 441.x, 443.1-443.9, 447.1, 557.1 557.9, V43.4 |
| Heart failure | I50.x | 398.91, 402.01, 402.11. 402.91, 404.01, 404.03, 404.11, 404.13, 404.91, 404.93, 425.4-425.9, 428.x |
| Renal failure | N18.x, N19.x | 403.01, 403.11, 403.91, 404.02, 404.03, 404.12, 404.13, 404.92, 404.93, 585.x, 586.x, 588.0, V42.0, V45.1, V56.x |
| Chronic liver disease | B18.x, B19.x, Z944, K9182, K70.x, K71.1, K71.3-K71.5, K71.7, K72.x-K74.x, K76.0, K76.2-K76.9. | 070.22, 070.23, 070.32, 070.33, 070.44, 070.54, 070.6, 070.9, 456.0-456.2, 570.x, 571.x, 572.2-572.8, 573.3, 573.4, 573.8,573.9, V42.7 |
| Chronic pulmonary disease | I27.8, 127.9, J40.x-J47.x, J60.x-J67.x, J68.4, J70.1, J70.3 | 416.8, 416.9, 490.x-505.x, 506.4, 508.1, 508.8 |
| Chronic Kidney Disease | I12.0, I13.1, N18.x, NI9.x, N25.0, Z49.0-Z49.2, Z94.0, Z199.2 | 403.01, 403.11, 403.91, 404.02, 404.03, 404.12, 404.13, 404.92, 404.93, 585.x, 586.x, 588.0, V42.0, V45.1, V56.x |
| Coagulopathy | D65-D68.x, D69.1, D69.3-D69.6 | 286.x, 287.1, 287.3-287.5 |
| Malignancy | C0x.x, C1x.x, C2x.x, C30.x, C31.x, C32.x, C33.x, C34.x, C37.x, C38.x, C39.x, C40.x, C41.x, C43.x, C45.x, C46.x, C47.x, C48.x, C49.x, C50, C51-58.x, C60-63.x, C76.x, C80.1, C81.x, C82.x, C83.x, C84.x, C85.x, C88.x, C9x.x | 196.x-199.x , 140.x-172.x, 174.x-195.x |
| Anemia | D50x, D51x, D52x, D53x, D630, D631, D638, D649 | 648.2, 285.2, 285.9, 280x, 281x, |
| Valve disease | A52.0, I05.x-I08.x, I09.1, I09.8, I34.x-I39.x, Q23.O- Q23.3, Z95.2, Z95.4 | 093.2, 394.x-397.x, 424.x, 746.3-746.6, V42.2, V43.3 |
| Hypothyroidism | E00.x-E03.x, E89.0 | 240.9, 243.x, 244.x, 246.1, 246.8 |
| **Previous history** | |  |
| Myocardial infarction | I25.2 | 412 |
| Stroke/TIA | Z86.73 | V1254, V1254, 438x |
| Medication non-compliance | Z91120, Z91128, Z91138, Z9114, Z9119 | V1581 |

|  | **ICD-10 CM/PCS codes** | **ICD-9 codes** |
| --- | --- | --- |
| **Thromboembolic events** | |  |
| Ischemic Stroke | I63.x | 434.x |
| Systemic embolic events | | |
| Upper and Lower Extremity embolism | I742, I744, I743 | 44421, 44422 |
| Other systemic embolism | I74.x | 444.x |
| TIA | G45.x | 435.x |

**Supplemental Table 2.** Trends in AF inpatients on documented anticoagulation therapy, stratified by CHADSVASC score in the United States.

|  | 2010 | 2011 | 2012 | 2013 | 2014 | 2015 | 2016 | 2017 | 2018 | 2019 | 2020 | 2021 | P Trend |
| --- | --- | --- | --- | --- | --- | --- | --- | --- | --- | --- | --- | --- | --- |
| **Long-term (current) anticoagulation** | | | | | | | | | | | | | |
| CHADSVASC 0 and 1 | 14.37 | 13.64 | 12.66 | 12.02 | 12.23 | 14.03 | 18.41 | 19.85 | 21.64 | 23.43 | 21.72 | 23.67 | <0.001 |
| CHADSVASC 2 to 4 | 158.46 | 172.82 | 166.68 | 168.37 | 176.74 | 205.02 | 252.12 | 280.27 | 309.89 | 340.14 | 321.63 | 348.68 | <0.001 |
| CHADSVASC ≥5 | 119.45 | 141.29 | 140.22 | 143.98 | 159.62 | 191.09 | 234.62 | 272.06 | 302.22 | 335.15 | 307.94 | 333.19 | <0.001 |
| Total | 292.29 | 327.75 | 319.56 | 324.37 | 348.59 | 410.14 | 505.15 | 572.18 | 633.75 | 698.72 | 651.29 | 705.54 | <0.001 |

**Supplemental Table 3.** Trends in AF inpatients undergoing percutaneous left atrial appendage occlusion procedures, stratified by CHADSVASC score in the United States.

|  | 2010 | 2011 | 2012 | 2013 | 2014 | 2015 | 2016 | 2017 | 2018 | 2019 | 2020 | 2021 | P Trend |
| --- | --- | --- | --- | --- | --- | --- | --- | --- | --- | --- | --- | --- | --- |
| **Percutaneous LAAO procedures** | | | | | | | | | | | | | |
| CHADSVASC 0 and 1 | 0.01 | 0.00 | 0.01 | 0.02 | 0.00 | 0.03 | 0.09 | 0.13 | 0.16 | 0.21 | 0.19 | 0.35 | <0.001 |
| CHADSVASC 2 to 4 | 0.11 | 0.08 | 0.07 | 0.17 | 0.13 | 0.39 | 1.36 | 2.53 | 3.73 | 5.52 | 5.65 | 9.91 | <0.001 |
| CHADSVASC ≥5 | 0.02 | 0.04 | 0.05 | 0.12 | 0.07 | 0.28 | 0.92 | 2.00 | 3.27 | 4.63 | 4.73 | 7.51 | <0.001 |
| Total | 0.14 | 0.12 | 0.13 | 0.31 | 0.20 | 0.71 | 2.38 | 4.66 | 7.16 | 10.36 | 10.58 | 17.78 | <0.001 |

**Supplemental Table 4.** Trends in crude hospitalization rates in ischemic stroke, transient ischemic attacks, and systemic embolisms stratified by CHADSVASC score in the United States.

|  | 2010 | 2011 | 2012 | 2013 | 2014 | 2015 | 2016 | 2017 | 2018 | 2019 | 2020 | 2021 | P Trend |
| --- | --- | --- | --- | --- | --- | --- | --- | --- | --- | --- | --- | --- | --- |
| **Ischemic stroke** | | | | | | | | | | | | | |
| CHADSVASC 0 and 1 | 1.33 | 1.42 | 1.41 | 1.35 | 1.56 | 1.59 | 1.85 | 1.81 | 1.90 | 1.85 | 1.63 | 1.83 | <0.001 |
| CHADSVASC 2 to 4 | 20.40 | 22.02 | 21.60 | 21.84 | 22.47 | 23.31 | 24.50 | 24.71 | 25.33 | 25.53 | 23.00 | 24.09 | 0.002 |
| CHADSVASC ≥5 | 15.77 | 17.68 | 18.20 | 18.54 | 19.46 | 21.11 | 22.34 | 23.78 | 23.58 | 24.33 | 21.56 | 22.27 | <0.001 |
| Total | 37.50 | 41.13 | 41.21 | 41.73 | 43.49 | 46.02 | 48.68 | 50.30 | 50.81 | 51.70 | 46.19 | 48.18 |  |
| **Transient ischemic attacks** | | | | | | | | | | | | | |
| CHADSVASC 0 and 1 | 0.37 | 0.33 | 0.35 | 0.39 | 0.29 | 0.28 | 0.26 | 0.25 | 0.22 | 0.17 | 0.13 | 0.16 | <0.001 |
| CHADSVASC 2 to 4 | 5.37 | 5.75 | 5.46 | 4.70 | 4.39 | 4.07 | 3.96 | 3.74 | 3.23 | 3.01 | 2.45 | 2.55 | <0.001 |
| CHADSVASC ≥5 | 4.61 | 5.30 | 5.05 | 4.77 | 4.34 | 4.15 | 4.10 | 4.15 | 3.98 | 3.60 | 2.86 | 3.11 | <0.001 |
| Total | 10.35 | 11.39 | 10.86 | 9.85 | 9.02 | 8.50 | 8.31 | 8.14 | 7.43 | 6.78 | 5.43 | 5.82 | <0.001 |
| **Systemic embolism** | | | | | | | | | | | | | |
| CHADSVASC 0 and 1 | 0.09 | 0.10 | 0.10 | 0.11 | 0.11 | 0.08 | 0.09 | 0.08 | 0.08 | 0.07 | 0.05 | 0.06 | 0.002 |
| CHADSVASC 2 to 4 | 1.47 | 1.45 | 1.50 | 1.42 | 1.48 | 1.33 | 1.19 | 1.15 | 0.95 | 0.95 | 0.76 | 0.73 | <0.001 |
| CHADSVASC ≥5 | 1.55 | 1.94 | 1.81 | 1.72 | 1.80 | 1.71 | 1.58 | 1.43 | 1.41 | 1.20 | 1.02 | 0.91 | <0.001 |
| Total | 3.11 | 3.49 | 3.40 | 3.26 | 3.38 | 3.12 | 2.87 | 2.65 | 2.45 | 2.21 | 1.82 | 1.70 | <0.001 |
| **All thromboembolic events** | | | | | | | | | | | | | |
| CHADSVASC 0 and 1 | 1.78 | 1.85 | 1.86 | 1.85 | 1.94 | 1.95 | 2.19 | 2.14 | 2.21 | 2.08 | 1.80 | 2.04 | 0.0690 |
| CHADSVASC 2 to 4 | 27.24 | 29.22 | 28.56 | 27.95 | 28.348 | 28.70 | 29.64 | 29.59 | 29.50 | 29.48 | 26.20 | 27.37 | 0.808 |
| CHADSVASC ≥5 | 21.92 | 24.92 | 25.04 | 25.03 | 25.59 | 26.97 | 28.01 | 29.35 | 28.96 | 29.11 | 25.43 | 26.28 | 0.022 |
| Total | 50.96 | 56.01 | 55.47 | 54.84 | 55.89 | 57.64 | 59.86 | 61.09 | 60.69 | 60.69 | 53.44 | 55.70 | 0.148 |

**Supplemental Table 5.** Trends in age-adjusted hospitalization rates in ischemic stroke, transient ischemic attacks, and systemic embolisms stratified by CHADSVASC score in the United States.

|  | 2010 | 2011 | 2012 | 2013 | 2014 | 2015 | 2016 | 2017 | 2018 | 2019 | 2020 | 2021 | P Trend |
| --- | --- | --- | --- | --- | --- | --- | --- | --- | --- | --- | --- | --- | --- |
| **Ischemic stroke** | | | | | | | | | | | | | |
| CHADSVASC 0 and 1 | 1.089 | 1.157 | 1.135 | 1.078 | 1.227 | 1.244 | 1.431 | 1.393 | 1.451 | 1.397 | 1.220 | 1.361 | 0.006 |
| CHADSVASC 2 to 4 | 16.755 | 17.901 | 17.376 | 17.399 | 17.737 | 18.228 | 18.978 | 18.982 | 19.307 | 19.325 | 17.216 | 17.951 | 0.095 |
| CHADSVASC ≥5 | 12.954 | 14.376 | 14.636 | 14.775 | 15.358 | 16.509 | 17.308 | 18.267 | 17.974 | 18.413 | 16.137 | 16.597 | 0.002 |
| Total | 30.80 | 33.43 | 33.15 | 33.25 | 34.32 | 35.98 | 37.72 | 38.64 | 38.73 | 39.13 | 34.57 | 35.91 | 0.009 |
| **Transient ischemic attacks** | | | | | | | | | | | | | |
| CHADSVASC 0 and 1 | 0.30 | 0.27 | 0.28 | 0.31 | 0.23 | 0.22 | 0.20 | 0.19 | 0.17 | 0.13 | 0.09 | 0.12 | <0.001 |
| CHADSVASC 2 to 4 | 4.41 | 4.68 | 4.40 | 3.74 | 3.47 | 3.18 | 3.07 | 2.87 | 2.46 | 2.28 | 1.83 | 1.90 | <0.001 |
| CHADSVASC ≥5 | 3.78 | 4.31 | 4.06 | 3.80 | 3.43 | 3.25 | 3.17 | 3.18 | 3.03 | 2.72 | 2.14 | 2.32 | <0.001 |
| Total | 8.50 | 9.26 | 8.74 | 7.85 | 7.12 | 6.65 | 6.44 | 6.25 | 5.66 | 5.13 | 4.07 | 4.34 | <0.001 |
| **Systemic embolism** | | | | | | | | | | | | | |
| CHADSVASC 0 and 1 | 0.07 | 0.08 | 0.08 | 0.09 | 0.08 | 0.06 | 0.07 | 0.06 | 0.06 | 0.05 | 0.03 | 0.04 | <0.001 |
| CHADSVASC 2 to 4 | 1.21 | 1.18 | 1.21 | 1.13 | 1.17 | 1.04 | 0.93 | 0.88 | 0.72 | 0.72 | 0.57 | 0.55 | <0.001 |
| CHADSVASC ≥5 | 1.27 | 1.58 | 1.45 | 1.37 | 1.42 | 1.34 | 1.23 | 1.10 | 1.08 | 0.91 | 0.76 | 0.68 | <0.001 |
| Total | 2.55 | 2.84 | 2.74 | 2.60 | 2.67 | 2.44 | 2.22 | 2.04 | 1.87 | 1.67 | 1.36 | 1.27 | <0.001 |
| **All thromboembolic events** | | | | | | | | | | | | | |
| CHADSVASC 0 and 1 | 1.47 | 1.51 | 1.50 | 1.48 | 1.54 | 1.53 | 1.70 | 1.64 | 1.69 | 1.58 | 1.35 | 1.52 | 0.608 |
| CHADSVASC 2 to 4 | 22.38 | 23.76 | 22.98 | 22.27 | 22.37 | 22.45 | 22.97 | 22.74 | 22.49 | 22.32 | 19.61 | 20.40 | 0.015 |
| CHADSVASC ≥5 | 18.01 | 20.26 | 20.15 | 19.95 | 20.20 | 21.09 | 21.71 | 22.55 | 22.08 | 22.04 | 19.04 | 19.59 | 0.265 |
| Total | 41.85 | 45.53 | 44.62 | 43.69 | 44.11 | 45.07 | 46.38 | 46.93 | 46.26 | 45.94 | 40.00 | 41.51 | 0.698 |
